# A statistical model of COVID-19 testing in populations: effects of sampling bias and testing errors

**DOI:** 10.1101/2021.05.22.21257643

**Authors:** Lucas Böttcher, Maria R. D’Orsogna, Tom Chou

## Abstract

We develop a statistical model for the testing of disease prevalence in a population. The model assumes a binary test result, positive or negative, but allows for biases in sample selection and both type I (false positive) and type II (false negative) testing errors. Our model also incorporates multiple test types and is able to distinguish between retesting and exclusion after testing. Our quantitative framework allows us to directly interpret testing results as a function of errors and biases. By applying our testing model to COVID-19 testing data and actual case data from specific jurisdictions, we are able to estimate and provide uncertainty quantification of indices that are crucial in a pandemic, such as disease prevalence and fatality ratios.

## 1. Introduction

Real-time estimation of the level of infection in a population is important for assessing the severity of an epidemic as well as for guiding mitigation strategies. However, inferring disease prevalence via patient testing is challenging due to testing inaccuracies, testing biases, and heterogeneous and dynamically evolving populations and severity of the disease.

There are two major classes of tests that are used to detect previous and current SARS-CoV-2 infections [1]. Serological, or antibody, tests measure the concentration of antibodies in infected and recovered individuals. Since antibodies are generated as a part of the adaptive immune system response, it takes time for detectable antibody concentrations to develop. Serological tests should thus not be used as the only method to detect acute SARS-CoV-2 infections. An alternative testing method is provided by viral-load or antigen tests, such as reverse transcription polymerase chain reaction (RT-PCR), enzyme-linked immunosorbent assay (ELISA), and rapid antigen tests, which are able to identify ongoing SARS-CoV-2 infections by directly detecting SARS-CoV-2 nucleic acid or antigen.

Test results are mainly reported as binary values (0 or 1, negative or positive) and often do not include further information such as the cycle threshold (Ct) for RT-PCR tests. The cycle threshold Ct defines the minimum number of PCR cycles at which amplified viral RNA becomes detectable. Large values of Ct indicate low viral loads in the specimen. An increase in Ct by a factor of about 3.3 corresponds to a viral load that is about one order of magnitude lower [2]. Cycle threshold cutoffs are not standardized across jurisdictions and range from values between 37 − 40, making it difficult to compare RT-PCR test results [3]. Lower Ct cutoffs in the range of 30 − 35 may be more reasonable to avoid classifying individuals with insignificant viral loads as positive [3].

Further uncertainty in COVID-19 test results arises from different type I errors (false positives) and type II errors (false negatives) that are associated with different assays. Note that inherent to any test, the threshold (such as Ct mentioned above) may be tunable. Therefore, besides intrinsic physical limitations, binary classification of “continuous-valued” readouts (*e.g*., viral load) may also lead to an overall error of either type [4]. In this work, we will assume that there is a standardized threshold and the test readout is binary; if any virus is detected, the test subject is positive. We will not explicitly model the underlying statistics of the errors but assume that the test readouts are binary but can be erroneous at specified rates. Some uninfected individuals will be wrongly classified as infected with rate FPR and some infected individuals will be wrongly classified as uninfected with rate FNR. For serological COVID-19 tests, the estimated proportions of false positives and false negatives are relatively low, with FPR ≈ 0.02 − 0.07 and FNR ≈ 0.02 − 0.16 [5–8]. The FNRs of RT-PCR tests depend strongly on the actual assay method [9,10] and may be significantly larger than those of serological tests. Typical values of FNR for RT-PCR tests lie between 0.1 and 0.3 [11,12] but might be as high as FNR ≈ 0.68 if throat swabs are used [7,12]. False-negative rates may also vary significantly depending on the time delay between initial infection and testing [8]. According to a systematic review [13] that was conducted worldwide, the initial value of FNR is about 0.54, underlying the importance of retesting. Similar to serological tests, reported false-positive rates of RT-PCR tests are about FPR = 0.05 [7].

Estimates of disease prevalence and other surveillance metrics [14,15] need to account for FPRs and FNRs, in particular if reported positive-testing rates [16] are in the few percent range and potentially dominated by type I errors. In addition to type-I/II testing errors, another confounding effect is biased testing [17], that is, preferential testing of individuals that are expected to carry a high viral load (*e.g*., symptomatic and hospitalized individuals). Biasing testing towards certain demographic and risk groups leads to additional errors in disease prevalence estimates that need to be corrected for.

To account for type-I/II errors, bias, retesting and exclusion after testing, we develop a corresponding framework for disease testing in Sec. 2. We apply our testing model to COVID-19 testing and case data in Sec. 3 and estimate testing bias by comparing random-sampling testing data [18] with officially reported, biased COVID-19 case data in Sec. 4. We conclude our study in Sec. 5.

## 2. Statistical testing model

Here, and in the following subsections, we develop a general statistical model for estimating the number of infected individuals in a jurisdiction by testing a sample population. The relevant variables and parameters to be used in our derivations are listed and defined in Table 1.

**Table 1.** Overview of variables used in testing model. An overview of the main variables and parameters that will be used in developing our testing model. The sets [0, *N*] and [0, *Q*] contain all integers from 0 up to *Q* and *N*, respectively. The set 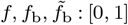 denotes all rational numbers between 0 and 1. For FNR, FPR, [0, 1] represents all real numbers between 0 and 1. We assume that 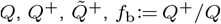 and 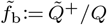 are determined by testing a population of known size *N*.

| symbol | definition |
| --- | --- |
| $N \in \mathbb{Z}^+$ | population in jurisdiction |
| $Q : [0, N]$ | number of tests administered |
| $Q^+ : [0, Q]$ | recorded positives under error-free testing |
| $\tilde{Q}^+ : [0, Q]$ | recorded positives under error-prone testing |
| $f : [0, 1]$ | true proportion of infected individuals |
| $f_b := Q^+ / Q : [0, 1]$ | fraction of positives under biased, error-free testing |
| $\tilde{f}_b := \tilde{Q}^+ / Q : [0, 1]$ | fraction of positives under biased, error-prone testing |
| $b \in \mathbb{R}$ | testing bias parameter |
| $\hat{b}, \hat{f} : [0, 1]$ | estimates of bias and underlying infection fraction |
| $\text{FPR} : [0, 1]$ | false positive rate |
| $\text{FNR} : [0, 1]$ | false negative rate |

Suppose we randomly administer *Q* tests within a particular short time period (*e.g*., within one day or one week) to a total effective population of *N* previously untested individuals. This population of individuals is comprised of *S* susceptible, *I* infected, and *R* removed (*i.e*., recovered or deceased) individuals, which are unknown. *S, I*, and *R* can dynamically change from one testing period to another due to transmission and recovery dynamics, as well as removal from the untested pool by virtue of being tested. The total population *N* = *S* + *I* + *R* can also change through intrinsic population dynamics (birth, death, and immigration), but can assumed to be constant over the typical time scale of an epidemic that does not cause mass death.

We start the derivation of our statistical model by first fixing *S, I* and *R*, assuming both perfect error-free testing, considering a “testing with replacement” scenario, in which tested individuals can be retested within the same time window. Under these conditions, the probability that *q* tests are returned positive and *Q*^−^ = *Q* − *q* tests are returned negative is

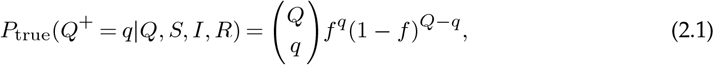

where the parameter

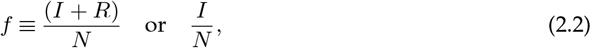

are simply the probabilities of identifying currently and previously infected individuals with tests such as serological (antibody) tests, or of detecting current infections with viral load tests, respectively. Note that testing with replacement renders *P*_true_ dependent only on *Q* and *f*, and not explicitly on *I, S, R* or *N*. The binomial expression 2.1 is accurate when the number of tests are much smaller than the population (*I* + *R*) or *I*.

Equation 2.1 describes perfect error-free and random testing. However, if there is some prior suspicion of being infected, the administration of testing may be biased. For example, certain jurisdictions focus testing primarily on hospitalized patients and people with significant symptoms [17], thus biasing the tests to those that are infected. We quantify such testing biases through a biased-testing function *B*(*Q*^+^) ∈ ℝ _≥0_, leading to the following modification of Eq. 2.1:

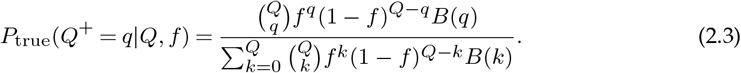

We define the biased-testing function *B*(*q*) as a weighting over certain numbers *q* of positive tests. A convenient choice of this weighting is a “Boltzmann” functional form 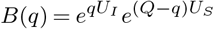, for which *P*_true_(*Q*^+^ = *q*|*f, Q*) becomes

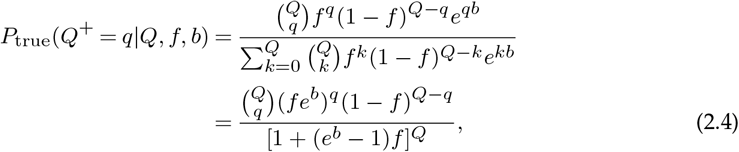

where −*U*_*I*_, −*U*_*S*_ represent “costs” for testing infecteds, susceptibles, and *b* ≡ *U*_*I*_ − *U*_*S*_ ∈ ℝ is a testing-bias parameter, which we employ below.

A Gaussian approximation to Eq. 2.4 can be found through the mean and variance of *Q*^+^:

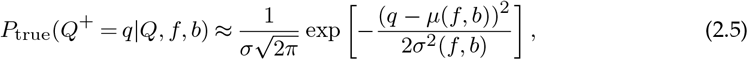

Where

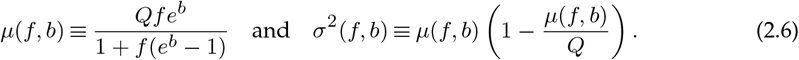

In addition to describing the probability distribution in Eq. 2.5 as a function of the number of positive tests *Q*^+^, we can also express it in terms of the observed positive (and potentially sample-biased) testing fraction *f*_b_ := *Q*^+^*/Q*:

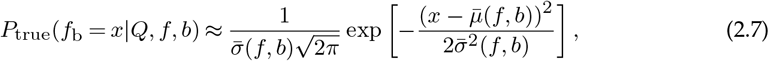

where here,

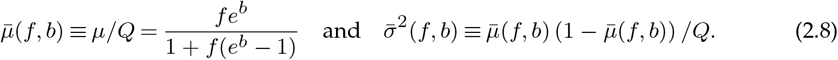

The expected value of 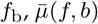, can be understood as a product of the true underlying infected fraction *f* and a bias function 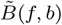 that depends on *f* and the bias parameter *b, i.e*., 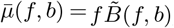, where 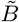 is given by

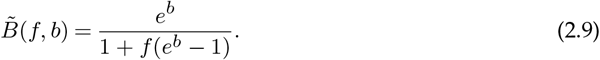

Note, that Eq. 2.9 implies that when *b >* 0, the currently and/or previously infected population is favored to be tested, while for *b <* 0, the non-infected and/or susceptible population is favored. The limits of *b* → ±∞ indicate testing that is completely biased such that only infected and susceptible individuals are tested, respectively. Realistic values of our bias parameter *b* are positive and ∼ *O*(1).

Fig. 1(a) shows the bias function 2.9 as a function of *b* for different infection fractions *f*. For 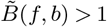, the biased-testing fraction 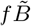 is larger than the unbiased-testing fraction *f*. The opposite holds for 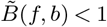. The variance 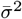 is plotted as a function of *b* in Fig. 1(b) and exhibits a maximum value of 1*/*(4*Q*) at *b*^∗^ = ln[(1 − *f*)*/f*].

**Figure 1.**
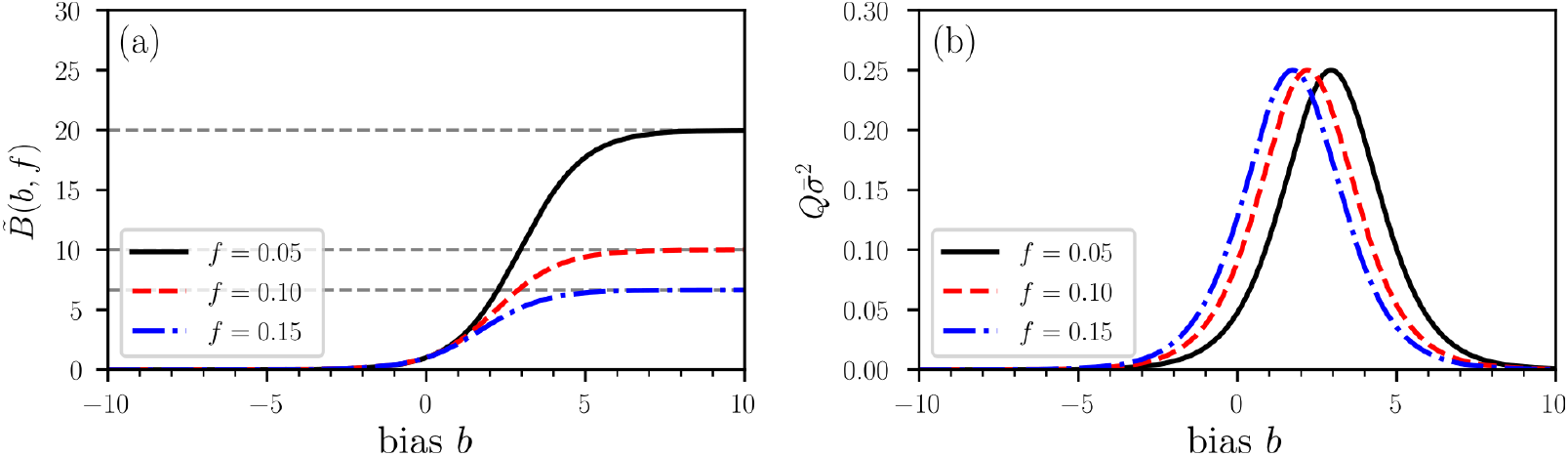
Illustration of a bias function 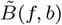. (a) The bias function 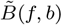 (Eq. 2.9) for three different fractions *f* of currently (and previously) infected individuals. Grey dashed lines indicate the asymptotic value 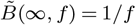. A value *b >* 0 indicates a testing bias towards currently and/or previously infected individuals while susceptible and/or non-infectious individuals are preferentially tested for *b <* 0. Unbiased testing corresponds to *b* = 0 and 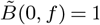 The variance 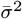 exhibits a maximum value of 1*/*(4*Q*) at a typical value of bias *b*^∗^ = ln[(1 *− f*)*/f*].

The probabilities *P*_true_ derived in Eqs. 2.1 and 2.3 correspond to “testing with replacement”. The opposite limit is “testing without replacement”; once an individual is tested they are labeled as such and removed from the pool of test targets, at least within the specified testing period. This concept of sampling with and without replacement commonly arises in the measurement of diversity in ecological settings [19]. Without replacement, and still under conditions of perfect random testing, two slightly different forms for *P*_true_ arise for the different type of tests (*e.g*., antibody *vs* PCR/viral load). For antibody tests that perfectly identifies recovered (or deceased) individuals as being previously infected, Eq. 2.1 is replaced by

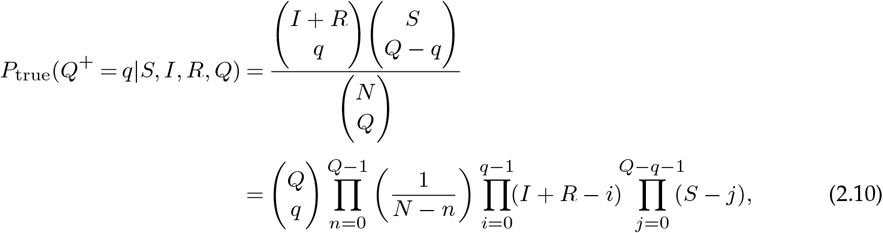

where the binomial coefficients for *q* = 0, *Q* are zero. On the other hand, if the perfect test only identifies individuals that currently have a viral load, the susceptible and recovered (or deceased) individuals both test negative and *P*_true_ is described by

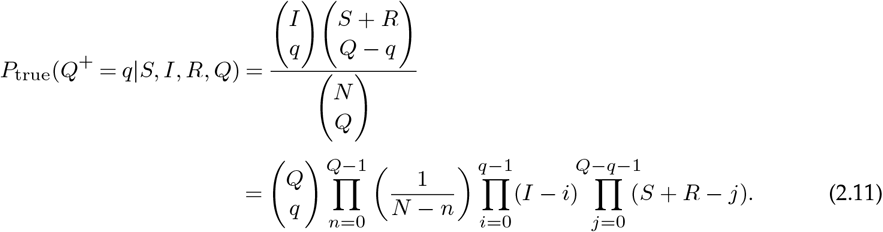

The expressions for *P*_true_ when tested subjects are not replaced, unlike in the case of testing with replacement, depend explicitly on *S, I, R*, and *N*.

To incorporate testing bias in into the probabilities *P*_true_ for testing without replacement, first consider Eqs. 2.11 and think of 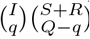 as the number of ways of distributing *q* p *I* infected individuals, and *Q* − *q* negative tests among *S* + *R* uninfected individuals. As in the biased-testing formulation of Eq. 2.3, we interpret the bias as a factor *B*(*q*) that weights more tests in *I* or *S* + *R* pools:

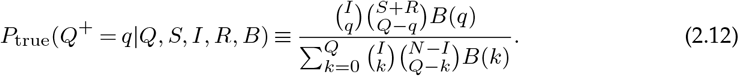

To obtain the “testing without replacement” equivalent of Eq. 2.4, we again set 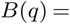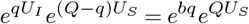. By using the Chu–Vandermonde identity

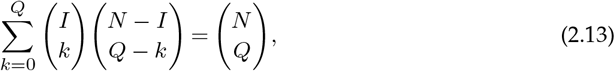

we verify that when *b* = 0 the expression 2.12 reduces to Eq. 2.11.

For the exponential form of *B*, the normalizing “partition function” becomes

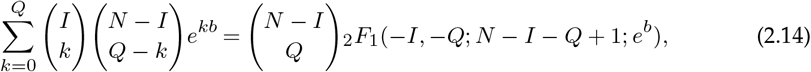

where _2_*F*_1_ denotes the (ordinary) hypergeometric function. Thus, the distribution of positive tests *Q*^+^ under biased testing without replacement for viral load-type tests can thus be expressed as

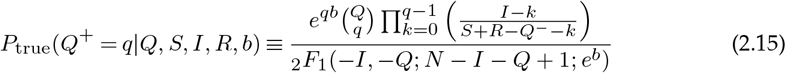

The distribution of positives under biased testing without replacement for antibody-type tests becomes (analogous to Eq. 2.10) is

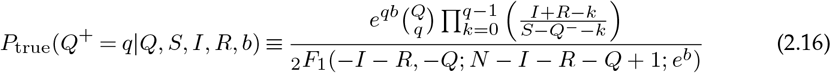

where *Q*^−^ ≡ *Q* − *q*. The above two expressions are equivalent except for the merging of the *R* pool with uninfected susceptibles *S* in one case, or with current infecteds *I* in the other. Because the testing decreases with the number of tests administered, Eqs. 2.15 and 2.16 cannot be reduced to functions of a simple positive-test fraction *f*_b_ or to universally accurate simple Gaussian forms.

### (a) Testing errors

The probability distributions *P*_true_ that we derived in Eq. 2.3 and in Eqs. 2.10–2.11 assume that testing is error-free, *i.e*., that the false-negative rate FNR = 1 − TPR = 0 and false-positive rate FPR = 1 − TNR = 0, or equivalently that the true-positive rate TPR = 1 and the true-negative rate TNR = 1. To incorporate erroneous testing, we now construct the probability distribution of error-generated deviation 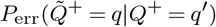 over the number of “apparent” positives *q* from tests that carry nonzero FPRs and FNRs, given that *q ′* positives would be recorded if the tests were perfect. If *q* apparent positive tests are tallied, *p* of them might have been true positives drawn from the perfect-test positives *q ′* in 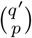 ways, while the remaining *k* = *q* − *p* apparent positives might have been erroneously counted as positives drawn from the *Q*^−^ ≡ *Q* − *q ′* true negatives. The remaining *q ′* − *p* true positive tests might have been erroneously tallied as false negatives, while the remaining *Q*^−^ − *k* negative tests might have been correctly tallied as true negatives. Assuming nonzero FPR and FNR, we find that the probability distribution of finding 0 ≤ *q* ≤ *Q* apparent positive tests is

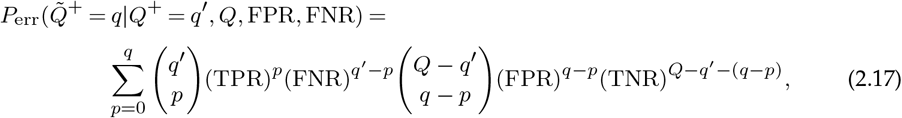

where we invoked the identities TPR + FNR = 1 and FPR + TNR = 1. The total error-prone distribution 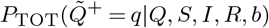 of recording *q* positives after having administered *Q* tests under bias and/or testing errors is given by convolving the probability *P*_err_ of finding *q* apparent tests given *q ′* true positive tests with the probability P_true_ of finding q ′ positive tests under perfect testing (Eq. 2.4 or Eq. 2.15):

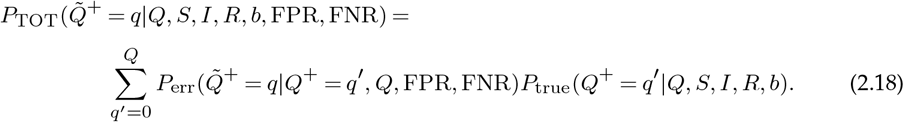

This convolution can be further simplified by taking the Gaussian limit of the binomial distributions that appear in *P*_err_ and in *P*_true_. To be concrete, we use *P*_true_ as written in Eq. 2.3 under the testing with replacement scenario and use the approximation 2.5. The same Gaussian approximation can be used for all terms in *P*_err_ from Eq. 2.17. After approximating the summation over *p* ∈ (0, *q*) in Eq. 2.17 by an integral over *p* ∈ (−∞, ∞) (provided *q* is sufficiently large), we find

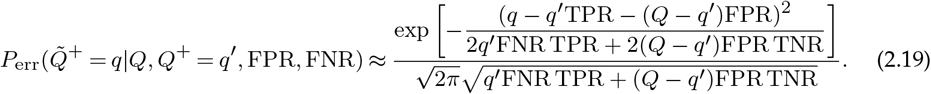

We now convolve Eq. 2.5 with Eq. 2.19 as prescribed by Eq. 2.18 to find

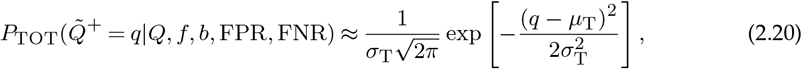

where

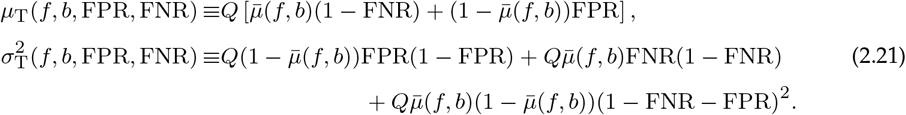

with 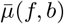 the mean value of the fraction of observed positives under biased, but perfect testing (see Eq. 2.8). The mean number of apparent positive tests *µ*_T_ is given by the sum of the expected value of true positive tests (*i.e*., 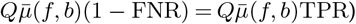 TPR) and the expected value of false positive tests (*i.e*., 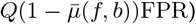 FPR). Based on the derived expressions for *µ*_T_ and 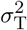, we define the random variable as the fraction of observed positive tests 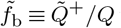 under biased and error-prone testing and obtain

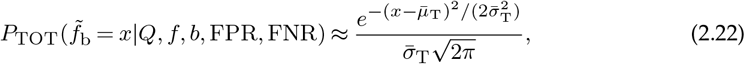

where

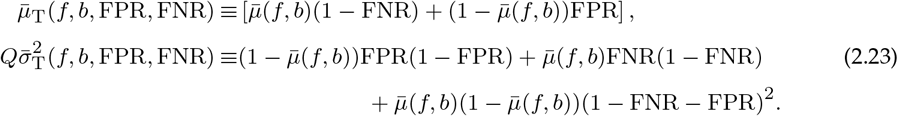

**Figure 2.**
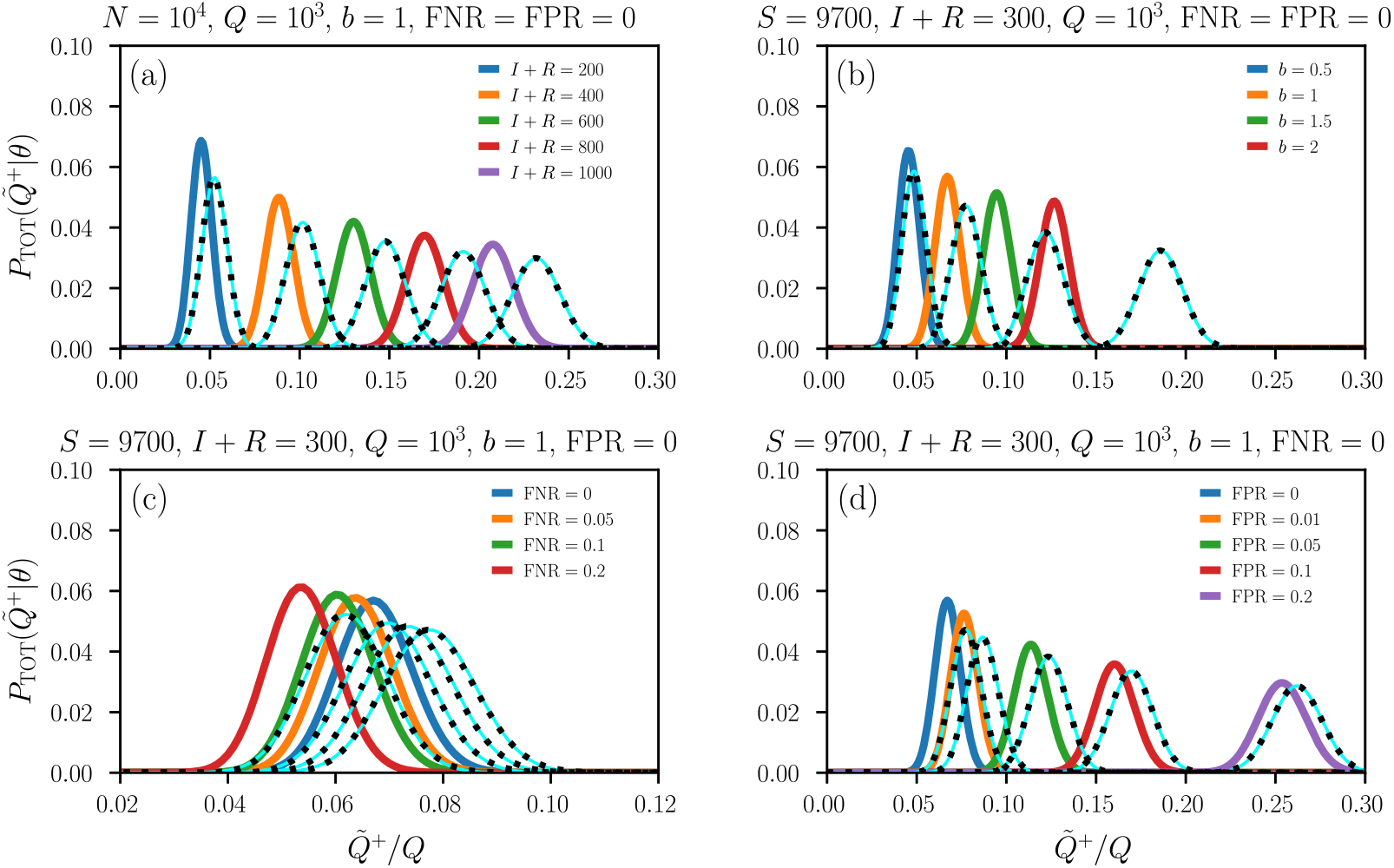
Distribution of apparently positive tests. Plots of 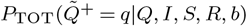 with *N* = *S* + *I* + *R* = 10^4^, *Q* = 10^3^, and different (a) values of *I* + *R*, (b) testing biases *b*, (c) FNRs, and (d) FPRs. The Gaussian approximation (solid light blue lines) of Eq. 2.22 provides an accurate approximation of *P*_TOT_. Dashed black lines correspond to distributions with replacement and the remaining solid colored lines correspond to those without replacement.

The Gaussian approximation 2.23 is quite accurate provided that (i) the number of positive and apparent positive tests, *Q*^+^ = *q ′* and 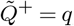, are sufficiently large, and (ii), the quantities 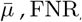, and FPR are not too close to 0 or 1. In Table 1, we provide an overview of the main variables used in the statistical testing model (2.17)–(2.23).

Fig. 2 shows the distribution of apparently infected individuals *P*_TOT_ for different numbers of infected/recovered individuals [Fig. 2 (a)], testing biases [Fig. 2 (b)], and testing sensitivities (*i.e*., true positive rates, TPRs) and specificities (*i.e*., true negative rates, TNRs) [Fig. 2 (c–d)]. Solid light blue lines represent the Gaussian approximation 2.22 and dashed black lines and the remaining colored lines are calculated by directly evaluating Eq. 2.18 with replacement (Eq. 2.3) and without replacement (Eq. 2.16), respectively. The FNRs that we consider in Fig. 2(c) are chosen in accordance with reported sensitivities of serological and RT-PCR tests for SARS-CoV-2 [9–12]. We observe that an increase in the FNR slightly shifts the distribution *P*_TOT_ towards smaller values of apparently infected individuals, which is consistent with the FNR dependence of the mean 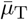 (Eq. 2.23). For serological and RT-PCR tests, the FPR = 1 − TNR is about 5%. A smaller specificity would lead to larger FPRs and a shift of *P*_TOT_ towards larger values of 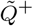 and 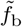 [Fig. 2(d)]. Our results show that *P*_TOT_ is more affected by variations in the testing specificity than by variations in testing sensitivity.

### (b) Temporal variations and test heterogeneity

Up to now, we have discussed single viral-load and antibody tests (with and without replacement) but have not considered temporal variations in the number of tests *Q*, the number of returned positives 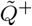, and heterogeneity in FNR and FPR that are associated with different classes (types, manufacturing batches, etc.) of assays. To make our model applicable to empirical time-varying testing data, we use *S*_*t*_, *I*_*t*_, *R*_*t*_ to denote the number of susceptible, infected, and removed individuals at time *t* (or in successive time windows labeled by *t*), respectively. If *K >* 1 test classes are present, we also include an additional index *c* ∈ {1, …, *K*} in all relevant model parameters. The testing bias and the total number of tests may be both test-class and time-dependent. That is, *b* = *b*_*t,c*_ and *Q* = *Q*_*t,c*_. Test specificity and sensitivity mainly depend on the assay type and not on time. We thus set FPR = FPR_*c*_ and FNR = FNR_*c*_.

## 3. Inference of prevalence and application to COVID-19 data

For a certain time window, one often wishes to infer *I*_*t*_ + *R*_*t*_ and *S*_*t*_, or *I*_*t*_ and *S*_*t*_ + *R*_*t*_ from values of *b*_*t,c*_, *Q*_*t,c*_, and *q*_*t,c*_. Alternatively, since *b*_*t,c*_ is difficult to independently ascertain, one may only be able to infer (*f*_*b*_)_*t,c*_ = *f* (*S*_*t*_, *I*_*t*_, *R*_*t*_, *b*_*t,c*_). For a single test result 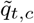 (or 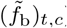), we can generate the maximum likelihood estimate (MLE) of the bias-modified prevalence 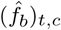 by setting the measured value 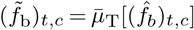 to find

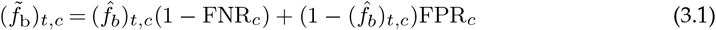

and

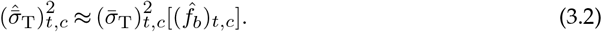

Since 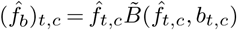, Eq. 3.1 can be solved for 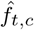:

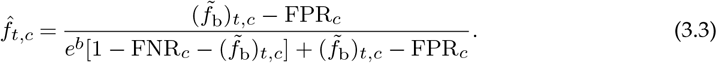

The posterior distribution *P*_post_ over values of *f* can be found through Bayes’ theorem:

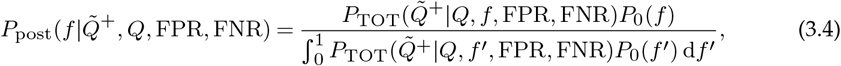

where *P*_0_(*f*) is a prior distribution over the underlying infection fraction *f* in the population. For notational brevity, we did not include the indices *c* and *t* in Eq. 3.4. We can again simplify the analysis by using the Gaussian approximation and a simple initial uniform prior, *P*_0_(*f < f*_max_ ≤ 1) = 1*/f*_max_.

As an example, we collected US testing data [20] from March 2020 to March 2021. Figure 3(a) shows the daily number of observed positive tests 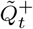 (red bars) and the corresponding total daily number of tests *Q*_*t*_ (blue bars). The 7-day average of the observed positive testing rate 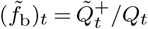 is indicated by the black solid line. The first drop in 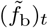 in March 2020 was associated with the initially very limited number of available SARS-CoV-2 testing infrastructure followed by the ramping up of testing capacity. After new cases surged by the end of March and in April 2020, different types of stay-at-home orders and distancing policies with different durations were implemented across the United States [21]. In June and July 2020, reopening plans were halted and reversed by various jurisdictions to limit the resurgence of COVID-19 [22].

**Figure 3.**
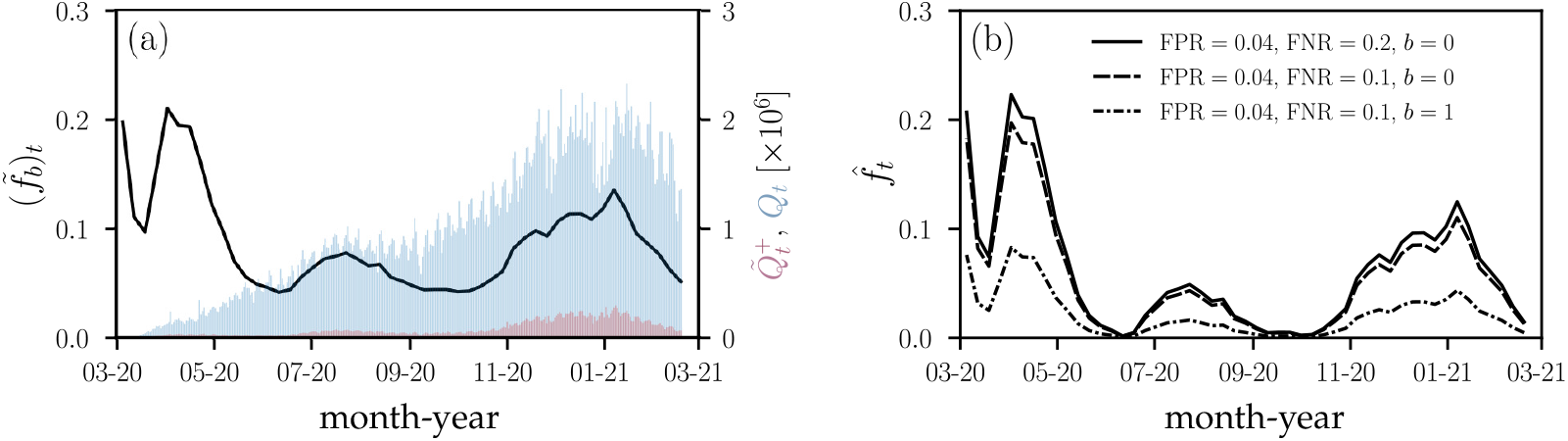
Observed and corrected proportions of positive tests in the United States. (a) The solid black line represents the 7-day average of the proportion of positive tests 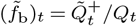 in the United States. Blue and red bars show the corresponding total number of daily tests *Q* and apparent positive tests 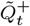, respectively. (b) The corrected proportion of positive tests 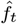, found by inverting Eq. 3.1, for different FPR, FNR, and bias combinations.

In Fig. 3(b), we show the corrected proportion of positive tests 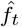, found by numerically inverting Eq. 3.1 for different FPR, FNR, and bias combinations. We observe that a small FPR = 0.04 shifts values 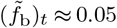 towards zero such that the corrected positive testing rate 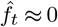. Reducing the FNR from 0.2 to 0.1 has only little effect on the corrected proportion of positive tests 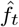 [solid black and dashed lines in Fig. 3(b)]. Accounting for a positive testing bias of *b* = 1 (*i.e*., preferential testing of infected and symptomatic individuals by a factor of *e*), however, markedly changes the inferred 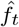 [dashed-dotted black line in Fig. 3(b)].

## 4. Inference of bias *b*

One way to estimate the testing bias *b* is to identify a smaller subset of control tests within a jurisdiction that is believed to be unbiased and compare it with the reported fraction of positive tests obtained via standard (potentially biased) testing procedures.

We can derive a rather complete methodology to estimate bias by formally comparing the statistics of two sets of tests applied to the same population. The first set of control tests with testing parameters *θ*_0_ = {*Q*_0_, *f*, FPR_0_, FNR_0_} is known to be unbiased (has prior distribution *δ*(*b*)), while the second set is taken with known parameters *θ* = {*Q*, FPR, FNR}, but unknown testing bias *b*. For example, the control set may consist of a smaller number *Q*_0_ of tests that are administered completely randomly, while the second set may be the scaled-up set of tests with *Q > Q*_0_. Since both sets of tests are applied roughly at the same time to the same overall population, the underlying positive fraction *f* is assumed to be the same in both test sets. We can then use Bayes’ rule on the first unbiased test set to infer *f* :

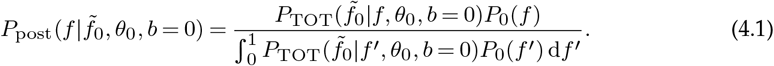

The probability distribution over *b* for a specified value of *f* can also be constructed from Bayes’ rule

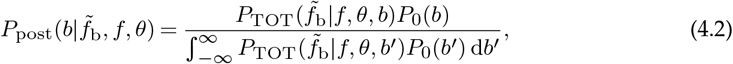

where *P*_0_(·) are prior distributions over the relevant parameters. The final distribution over the bias factor, given the two measurements 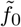 and 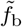 derived from the two sets of tests with testing parameters *θ*_0_ and *θ*, can be found using

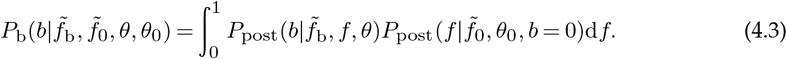

Of course, a simpler maximum likelihood estimate can also be applied to data by first inferring the most likely value of *f* from the control test set. We can use the number of positive tests in the control sample 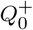 to define the variable 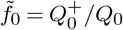. One can then maximize 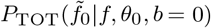 with respect to *f* and use this value 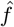 in 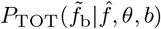. Maximizing 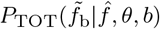 with respect to *b* then gives the MLE estimate 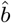. We can use random and unbiased sampling results obtained in the German jurisdiction of Gangelt, North Rhine-Westphalia [18]. A total of 600 adult persons with different last names were randomly selected from a population of 12,597 and asked to participate in the study together with their household members. The resulting study comprised of *Q*_0_ = 919 subjects who underwent serological and PCR testing between March 31-April 6, 2020. The specificity and sensitivity corrected, unbiased positive test fraction was determined to be *f* = 15.53% (95% CI; 12.31%–18.96%). Thus, we use this value as an estimate for the true underlying positivity rate 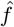. The larger sample taken across North Rhine-Westphalia between March 30–April 5, 2020 was measured (*Q* ≈ 25, 000) to be 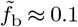 [23]. Assuming that this value is also error-corrected, an estimate of the bias 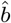 in this main testing set can be found by solving 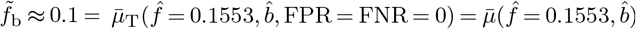 for 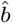. We find that the difference between the unbiased positive testing rate of 15.53% and 10% corresponds to a bias of 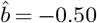. This negative bias likely arises because Gangelt was a an infection hotspot within the entire North Rhine-Westphalia region, so the control sample was probably not unbiased. For comparison, a higher biased positive testing rate of 20% would lead to an estimated testing bias 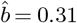.

The number of total deaths on April 6, 2020 amounted to 7. Hence, the corresponding estimate of the infection fatality ratio (IFR), the number of disease-induced deaths *D*_*t*_ divided by the total number of cases *N*_*t*_ at time *t*, in this jurisdiction on April 6, 2020 was 7*/*(0.1553 × 12,597) = 0.36% (95% CI; 0.29%–0.45%) [18]. If only a biased estimate of the proportion of positive cases is known and not the true value *f*, we can use our framework to distinguish between the true IFR_*t*_ = *D*_*t*_*/*(*N*_*t*_ − *S*_*t*_) = *D*_*t*_*/*(*f*_0_*N*_*t*_) and the observed infection fatality ratio

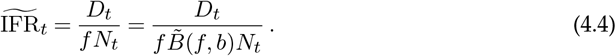

Figure 4 shows the observed 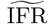 as a function of testing bias *b* for the aforementioned example of the German jurisdiction of Gangelt. Values of *b >* 0 correspond to preferential testing of infected individuals and thus lead to an apparently lower 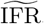. The opposite holds for *b <* 0.

**Figure 4.**
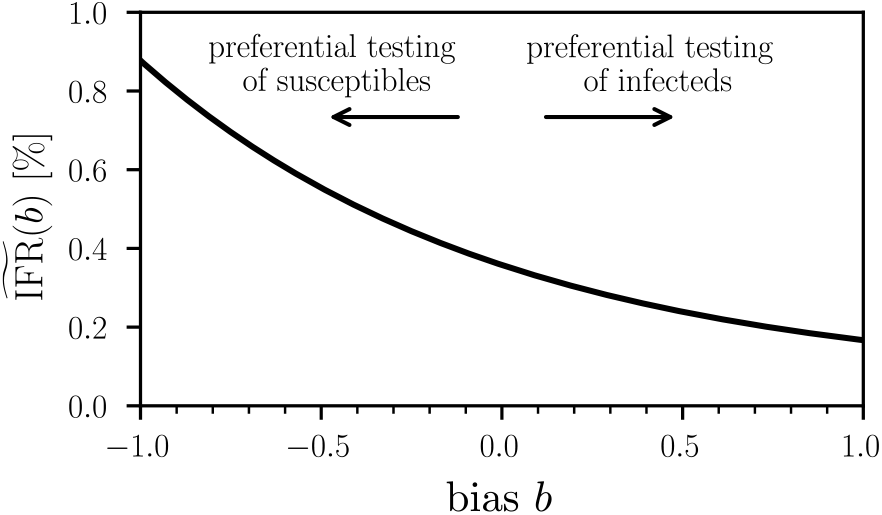
Dependence of the observed IFR on testing bias. The observed infection fatality ratio 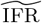 (Eq. (4.4)) as a function of testing bias *b*. We used the example of the German jurisdiction Gangelt and set 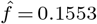, *D* = 7, and *N* = 12,597. A value *b >* 0 indicates a testing bias towards currently and/or previously infected individuals while susceptible and/or non-infectious individuals are preferentially tested for *b <* 0. Unbiased testing corresponds to *b* = 0.

## 5. Summary and Conclusions

Radiological testing methods such as chest computed tomography (CT) are used sporadically to identify COVID-19-induced pneumonia in patients with negative tests [24], However, the overwhelming majority of COVID-19 tests are based on serological (or antibody) tests and rapid antigen tests, ELISA, and RT-PCR assay [1]. These tests are designed to subsequently output a binary signal, either infected or not. The population statistics of this output are affected by testing errors and bias. False positive and false negative rates of serological tests are generally smaller than those of rapid antigen tests and RT-PCR tests. However, serological tests are unable to identify early-stage infections since they are measuring antibody titres that usually develop a few days up to a few weeks after infection. In addition to the occurrence of false positives and false negatives (*i.e*., type-I and type-II errors), certain demographic groups (*e.g*., elderly people or those with comorbidities such as heart and lung diseases) may be overrepresented in testing statistics.

To quantify the impact of both type-I/II errors and testing bias on reported COVID-19 case and death numbers, we developed a mathematical framework that describes erroneous and biased sampling (both with and without replacement) from a population of susceptible, infected, and removed (*i.e*., recovered or deceased) individuals. We identify a positive testing bias *b >* 0 with an overrepresentation of previously or currently infected individuals in the study population. Conversely, a negative testing bias *b <* 0 corresponds to an overrepresentation of susceptible and/or non-infectious individuals in the study population. We derived maximum likelihood estimates of the testing-error and testing-bias-corrected fraction of positive tests. Our methods can be also applied to infer the full distribution of corrected positive testing rates over time and for different types of tests across different jurisdictions.

The mathematical quantity that underlies most of our analysis is the proportion of apparent positive tests. As pointed out in [25], the *absolute* number of positive tests may not capture the actual growth of an epidemic due to limitations in testing capacity. Still, many jurisdictions report absolute case numbers without specifying the total number of tests or additional information about test type, date of test, and duplicate tests [26], rendering interpretation and application to epidemic surveillance challenging. For a reliable picture of COVID-19 case numbers, more complete testing data, including total number of tests, number of positive tests, test type, and date of test, has to be reported and made publicly available at online data repositories. To correct for false positive, false negatives, and testing bias in testing statistics (Fig. 3), it will be also important to further improve estimates of FPR, FNR, and *b* through in field studies. In particular, estimating the testing bias *b* requires random sampling studies similar to that carried out in [18]. Finally, while we have presented our analysis in the context of the COVID-19 pandemic, the general results presented in this paper apply to testing and estimation of severity of any infectious disease afflicting a population.

## Data Availability

Our source codes are publicly available at https://github.com/lubo93/disease-testing.

## Data Accessibility

Our source codes are publicly available at https://github.com/lubo93/disease-testing.

## Authors’ Contributions

LB, MRD, and TC contributed equally to the study design, data analyses, and manuscript writing.

